# Illness and Wage Loss: Longitudinal evidence from India (and Implications for the Universal Health Coverage Agenda)

**DOI:** 10.1101/2021.11.04.21265892

**Authors:** Aditya Srinivas, Suhani Jalota, Aprajit Mahajan, Grant Miller

## Abstract

**Background:** A key aim of Universal Health Coverage (UHC) is to protect individuals and households against the financial risk of illness. Large-scale health insurance expansions are therefore a central focus of the UHC agenda. Importantly, however, health insurance does not protect against a key dimension of financial risk associated with illness: forgone wage income (due to short-term disability). In this paper, we quantify the economic burden of illness in India attributable – separately – to wage loss and to medical care spending, as well as differences in them across the socio-economic distribution.

**Methods:** We use data from two Indian longitudinal household surveys: (i) the Village Dynamics in South Asia (VDSA) survey (1,350 households surveyed every month for 60 months between 2010 and 2015) and (ii) the Indian Human Development Survey (IHDS) (more than 40,000 households surveyed in 2005 and again in 2011). The VDSA allows us to study the economic dynamics of illness using high-frequency observations, and the IHDS allows us to confirm our findings in a nationally-representative sample. Both contain individual- and household-level information about illness, wage income, and medical spending over time. We use longitudinal variation in illness to estimate regression models of economic burden separately for wage loss and medical care spending across the socio-economic distribution. Our regression models include a series of fixed effects that control for differences in time-invariant household (or individual) characteristics and time-varying factors common across households.

**Findings:** 1,184 households (88%) in the VDSA sample reported an episode of illness over 60 months, and 15770 households (40%) in the IHDS reported an illness in the preceding year. In the VDSA sample, on average, a day of illness was associated with a reduction in monthly per capita wage income of Rs 77 [95% CI −99 to −57] and an increase in monthly per capita medical spending of Rs 126 [95% CI 110-142]. Variation across the socio-economic distribution was substantial. Among the poorest households, wage loss due to illness is roughly 15% of total household spending – nearly three times greater than medical spending. Alternatively, among the most affluent households, wage loss is less than 5% of total household spending – and only one-third of medical spending. Put differently, wage loss accounts for more than 80% of the total economic burden of illness among the poorest households, but only about 20% of the economic burden of illness among the most affluent. Estimates from the IHDS sample show that this socio-economic gradient is present in the Indian population generally.

**Interpretation:** Wage loss accounts for a substantial share of the total economic burden of illness in India – and disproportionately so among the poorest households. If Universal Health Coverage truly aims to protect households against the financial risk of illness – particularly poor households, the inclusion of wage loss insurance or another illness-related income replacement benefit is needed.

Research in context

Evidence before this study
We searched Google Scholar for the terms “burden of illness, economic impact of health shock, poor households” and found several systematic reviews of the economic burden of illness in low- and middle-income countries. We then reviewed these articles and the ones that they incorporate. The economic burden of illness among households in low- and middle-income countries is well-established and substantial. However, the vast majority of existing studies focus only on economic burden due to medical care spending, with limited and mixed evidence on wage loss. One systematic review concludes that the economic burden of illness on wage loss is unclear, with some studies showing no effect at all.

Added value of this study
Our paper makes two contributions. First, it directly estimates and compares the magnitude of wage loss and direct medical care expenses due to illness using detailed, high frequency data on wage earnings and illness. Past studies have largely focused on either total economic burden or medical care costs, but our direct comparison informs health policies focused on strengthening financial protection. Our high frequency data allow us to pinpoint precise temporal relationships between illness and its economic burden that have not been possible with other commonly-used longitudinal surveys.
Second, given the large number of longitudinal observations for each household in the data, our paper is the first to produce *household-specific* estimates of the burden of illness, enabling us to trace-out both wage loss and medical care expenses flexibly across the distribution of household economic status - enabling a more granular analysis of disparities between the poor and more affluent. We show that in India, which accounts for nearly half of those impoverished due to illness globally, wage loss accounts for more than 80% of the total economic burden of illness among the poorest households, but only about 20% of the economic burden of illness among the most affluent ones.

Implications of the available evidence
In contributing to existing evidence, the findings of this paper suggest that even if successful, the current Universal Health Coverage (UHC) agenda may still not protect poor households in India (and potentially many low- and middle-income countries) from substantial economic burden due to illness. If Universal Health Coverage in countries like India truly aims to protect households against the total economic burden of illness – particularly poor households, wage loss insurance or another health-related income-replacement benefit is required along with health insurance.

## Introduction

A central aim of Universal Health Coverage (UHC) is to protect individuals and households against the financial risk of illness – with particular emphasis on poor, vulnerable, and marginalized segments of the population.^1^ This goal is supported by a substantial body of evidence estimating that nearly 100 million people worldwide are pushed into extreme poverty each year because of out-of-pocket spending for unanticipated medical care needs.^2^ The rise in non-communicable diseases (NCDs) is pushing these estimates higher,^3^ and the SARS-CoV-2 pandemic is presumably as well. Globally, India – the focus of this paper – accounts for nearly half of those impoverished due to illness, and illness is a leading cause of indebtedness and poverty.^4–8^ Large-scale health insurance expansions in India, and in low-and middle-income countries generally, are therefore a central focus of the UHC agenda.^9–13^

Importantly, however, health insurance does not protect against the other key dimension of financial risk associated with illness: wage loss. In many high-income countries, health and wage loss (or short-term disability) insurance exist in tandem to protect against both forms of risk.^14–17^ However, in lower-income countries, wage loss insurance or illness-related income replacement benefits are often absent, meaning that for every day a working individual is sick and unable to work, that individual is not paid (and coverage of existing programs often excludes informal workers).^18^ This concern that applies disproportionately to the poor, who are more likely to be day-laborers (without sick leave) rather than salaried employees.

Nonetheless, the extent to which wage loss due to illness is an important source of economic burden in lower-income countries – and its magnitude relative to out-of-pocket medical spending – is largely unmeasured. A large body of research documents a substantial economic burden of illness among households in India and in low- and middle-income countries generally.^19–22^ However, this research has largely focused on out-of-pocket medical spending,^23–27^ with limited evidence on wage loss.^28–31^ One systematic review concludes that the economic burden of illness due to wage loss is unclear, with some studies suggesting no effect at all.^18^ Moreover, few studies have explicitly considered the relative size of wage loss and out-of-pocket medical spending (the two major components of financial risk associated with illness)^30–34^ – nor differences in them across the socio-economic distribution.^35^

In this paper, we quantity the relative importance of wage loss and medical spending due to illness, as well as variation in their relative magnitude across the socio-economic distribution. We also demonstrate that although standard measures of financial risk due to illness (defined as household medical spending as a share of total spending) sometimes counterintuitively suggest that more affluent households face relatively greater risk (presumably due, in part, to inequality in access to health care), this result is reversed when wage loss is incorporated.^36,37^ Ultimately, our results demonstrate the significance of disability risk (or wage loss due to illness) as an important but under-recognized issue relevant to the UHC agenda – particularly among the poorest households.

## Methods

### Data Sources

We use two longitudinal datasets to estimate wage loss and medical spending due to illness among households across the socio-economic distribution: the Village Dynamics in South Asia (VDSA) survey, collected by the International Crops Research Institute for the Semi-Arid Tropics (ICRISAT), and the Indian Human Development Survey (IHDS).

The ICRISAT VDSA survey (hereafter VDSA) is a household longitudinal survey of 1,400 households conducted every month for 60 consecutive months between July 2010 and June 2015. Households were randomly selected from 30 villages in eight Indian states (Andhra Pradesh, Bihar, Gujarat, Jharkhand, Karnataka, Madhya Pradesh, Maharashtra, and Orissa). Four villages were selected from each state (except Madhya Pradesh, from which two were selected) to represent the agro-climatic conditions in India’s semi-arid and humid tropical regions, as shown in Appendix Figure A1. Households in each village were in turn randomly selected to represent households in four landholding classes: large, medium, small, and landless. The survey collected monthly data on labor force participation (number of days worked and wage income) and self-reported illness for each member of the household, as well as consumption and income for the household.

For our purposes, the high frequency collection of data on illness, wages, and spending at the household level is particularly useful (and rare). For each household in the VDSA sample, we code an illness episode for a household as a binary outcome equal to 1 if any member in the household reported an illness in a given month, and zero otherwise. We also code wage income for households as the sum of earned wage income for all household members (capturing compensating changes in the labor supply of other household members when an individual becomes sick) and income earned from household businesses. To measure medical care spending, we use monthly data on total household spending for medical care.

Despite the VDSA survey’s detail and high frequency, it is not representative of the broader Indian population. Therefore, in harmonized parallel analyses, we also use the VDSA survey together with data from the Indian Human Development Survey (IHDS) to consider the extent to which our VDSA results generalize to the rest of India. The IHDS is a nationally representative longitudinal household survey fielded in 2005 and again in 2011, collecting data from 42,152 households in 1,503 villages and 971 urban neighborhoods across all major Indian states. The IHDS contains comprehensive information about major illnesses over the past year for each individual in the household (major illnesses include diagnoses of cataracts, tuberculosis, hypertension, and heart disease), the corresponding medical spending over the past year for each episode of illness, and total household income by source over the past year.

To conduct harmonized analyses that can be implemented in both the VDSA and IHDS samples in the same way (with comparable measurement of illness, wage income, and medical spending over the same lengths of time), we construct individual-year observations in each dataset. We code illness episodes in both datasets as an individual’s number of sick days in the preceding year due to major illness. To measure household wage income, we sum monthly data on wage income in the VDSA data, and we use wages exclusive of benefits or bonuses in the IHDS data. For medical spending, we sum monthly data on medical spending in the VDSA data, and we use total spending for services, medicines, hospitalizations, and travel to health facilities in the IHDS data.

Finally, to measure the socio-economic status of households in both surveys at the start of their survey periods, we follow a standard convention in economics by using baseline total household spending per capita (henceforth “economic status” – consumption spending per capita, broadly defined, is a widely-used measure of economic well-being^38–40^ and has been used by others studying the effect of illness on consumption spending^30^). Appendix Table A1 provides summary statistics for both the VDSA and IHDS samples.

### Statistical Analysis

Our statistical approach uses longitudinal variation within households in both the VDSA and IHDS samples to estimate the economic burden of illness separately for wage income and medical spending. Because the structure (and frequency) of two data sources differ, we use variants of the same general statistical framework, focusing first on the VDSA data (to capitalize on the strengths of its monthly survey waves), and then conducting parallel analyses using a statistical framework that can be implemented in the same way in both datasets (to consider how our results generalize to the broader Indian population).

Our statistical analysis proceeds in two steps. We first leverage the richness of the VDSA sample (with 60 monthly observations per household) to estimate the following household-level model:

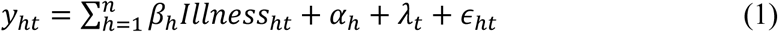

where *y*_*ht*_ is an outcome for household *h* in month *t* (either wage income per capita or medical spending per capita); *illness*_*ht*_ is an indicator variable equal to 1 if any member in household *h* reported an illness in month *t* and 0 otherwise; and *α*_*h*_ and *λ*_*t*_ are household and month fixed effects, respectively. This approach uses within-household variation in illness episodes over time to estimate the *household-specific* burden of illness separately for wage income and medical spending (i.e. the coefficient on illness differs across households). The remarkably high frequency VDSA sample enables us to study the household-specific temporal nuances of illness episodes and their consequences. Importantly, because the two components of total economic burden may vary considerably between poorer and wealthier households, we use household-specific estimates and show that the relative magnitudes of wage loss and medical spending vary systematically across the distribution of socio-economic status. We plot these household-specific estimates by economic status using a flexible polynomial fit.

Then, using a regression framework that can be implemented in the same way in both the VDSA and IHDS samples, we estimate the economic burden due to wage loss and medical spending at the individual-year level:

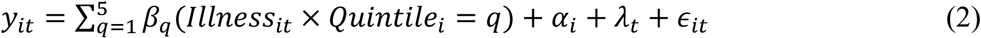

Where *illness*_*it*_ is the number of days of illness in year *t* for individual *Q, Quintile*_*i*_ is individual *i*’s economic quintile (of the baseline per capita expenditure distribution); and *α*_*i*_ and *λ*_*t*_ are individual and year fixed effects, respectively. This approach compares the wage income and medical spending of individuals that experienced an illness to those that did not.

We control for a series of fixed effects in both of our statistical models. The household fixed effects *α*_*h*_ (or individual fixed effects *α*_*i*_) control for differences in time-invariant household (or individual) characteristics such as wealth or long-term health, while the time fixed effects *λ*_*t*_ control for time-varying events common to all households such as economic downturns. We used STATA statistical software for all analyses, and our statistical code is available upon request.

## Results

### VDSA Sample

In the VDSA sample, over a period of 60 months, 1,184 households (88%) reported an episode of illness, 1,337 households (99%) reported medical care spending, and 1,270 households (95%) reported participating in the labor market. Average monthly wage income was Rs. 1169 per capita per month, average medical spending was Rs. 93 per capita per month, and total household spending was Rs. 1160 per capita per month.

Figure 1, Panel A graphs household-specific estimates of the economic burden of illness from Equation (1), separately for wage income and medical spending, across the distribution of household economic status (measured as the logarithm of baseline household spending per capita) – ranging from poorest households (on the left, with a log value of 6, equivalent to household spending of Rs 400 per capita or 8·75 USD) to most affluent households (on the right, with a log value of 8, equivalent to a household spending of Rs 2900 per capita or 63·5 USD). To smooth the gradient of these relationships across the economic distribution, we use a fractional polynomial fit. For each household-specific estimate, the figure also shows 95% confidence intervals. For a household at the mean of the economic distribution, a day of illness is associated with a reduction in monthly wages of Rs 53 [95% CI −10 to −70] per capita and an increase in medical spending of Rs 72 [95% CI 50–95] per capita. Appendix Table A2 also reports regression estimates of the average per capita burden of illness due both to wage loss and medical care spending (Rs 77 [95% CI −99 to −57] and Rs 126 [95% CI 110–142], respectively).

**Figure 1:**
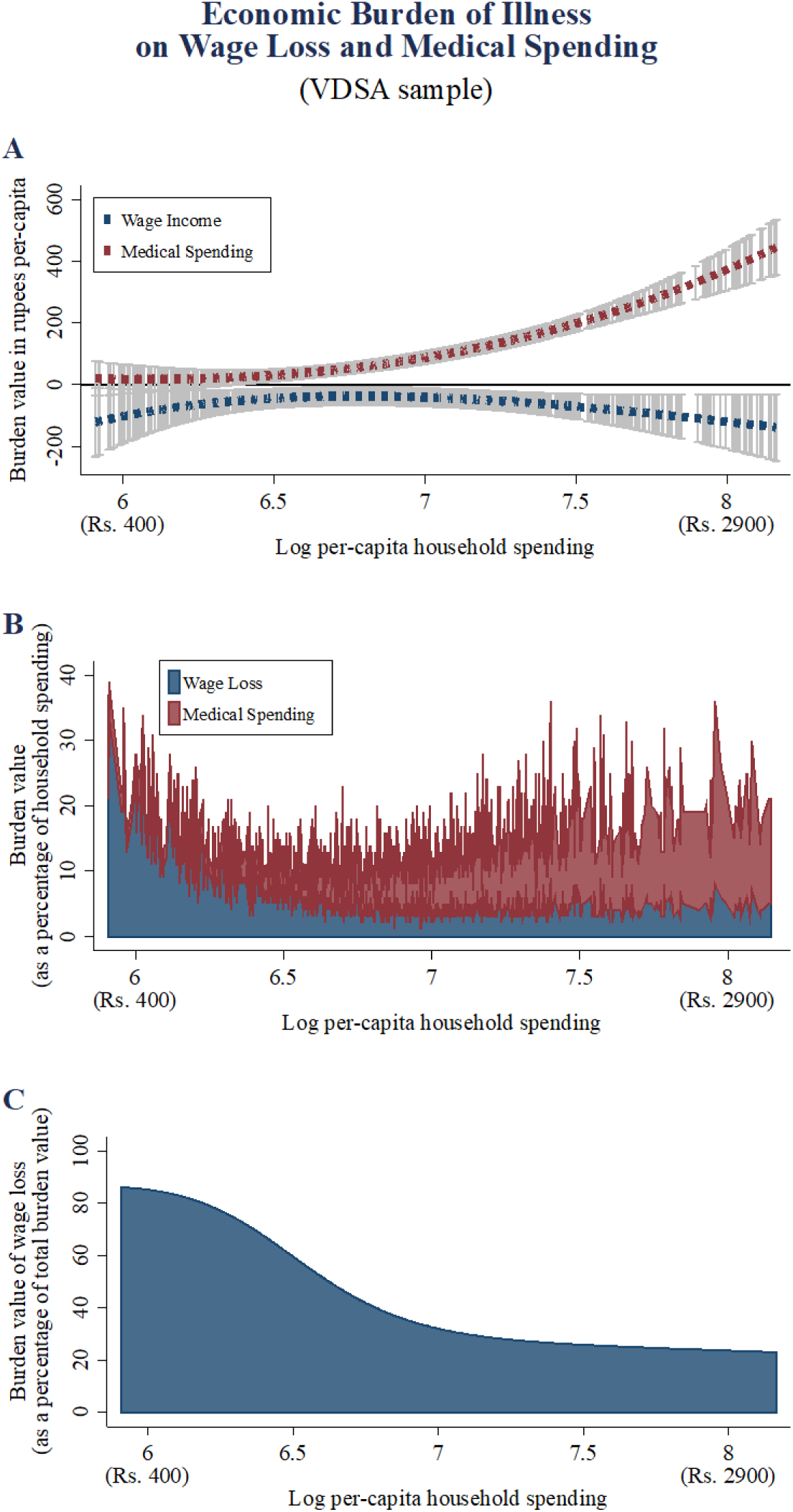
Panels A-C use VDSA sample data of 1300 households collected monthly between July 2010 and June 2015. Panel A plots fitted values from a flexible polynomial regression of the estimates from Equation (1) on the logarithm of 2010 per-capita household spending. The regression includes a second-degree fractional polynomial of logarithm of 2010 per-capita spending as a covariate. The grey bars represent 95% confidence intervals of the flexible fractional polynomial fit. A logarithm value of 6 (on the left) is equivalent to household spending of Rs 400 per capita or 8·75 USD, and a logarithm value of 8 (on the right) is equivalent to household spending of Rs 2900 per capita or 63·5 USD. Panel B decomposes the total economic burden of illness into its two components – wage loss (in blue) and medical expenses (in red), and shows the economic burden (for both wage loss and medical expenses) as a percentage of total household spending. Finally, Panel C displays the proportion of the total economic burden of illness that is attributable to lost wages.

Considering how these effects vary across the income distribution, the absolute value of wage loss due to illness is greater among poorer households, reaching nearly Rs. 150 per capita per month [95% CI 50–200] among the poorest (relative to median baseline household spending of Rs. 550 per capita per month). This relationship is then generally flat across the middle of the distribution of household economic status, increasing again in absolute magnitude among the most affluent households (at the right), reaching Rs. 150 per capita per month [95% CI 20–220] (relative to median baseline household spending of Rs. 2000 per capita per month). Alternatively, medical spending due to illness is relatively low among poorer households (below Rs. 50 per capita per month [95% CI −10 – 50]), and then rises steeply among wealthier ones, reaching Rs. 450 per capita per month [95% CI 350–500] (presumably reflecting, at least in part, underlying differences in access to medical care).

Figure 1, Panel B explicitly decomposes the two major components of total economic burden due to illness across the economic distribution, with the burden value from medical spending (in red) stacked on top of the burden value from wage loss (in blue). To examine more directly how the relative magnitudes shown in Panel A compare to baseline household spending, Figure 1 Panel B shows, at each point across the economic distribution, the relative size of wage loss and medical spending as a share of total household spending at baseline. Strikingly, among the poorest households, wage loss due to illness is around 15% of total household spending – nearly three times greater than the size of medical spending (which is approximately 5% of household spending). Alternatively, for the most affluent households, wage loss is less than 5% of total household spending, while medical spending is about three times greater, at roughly 15% of total spending.

Figure 1, Panel C then summarizes these results, showing the share of the total burden of illness (measured as the sum of wage loss and medical spending) due to lost earnings. For the poorest households, wage loss accounts for more than 80% of the total economic burden of illness, but only about 20% of the burden among the most affluent households.

### Harmonized VDSA and IHDS Samples

We then use the harmonized VDSA and nationally-representative IHDS samples to consider the extent to which the VDSA results (shown in Figure 1) generalize to the broader Indian population. As Appendix Table A1 shows, in the IHDS sample, more than 15770 households (40%) reported an illness in the preceding year. Average annual wage income is Rs. 12,201 per capita, average annual medical spending is Rs. 1,270 per capita, and average total annual household spending is Rs. 22,737 per capita. The summary statistics for the harmonized VDSA sample are quantitatively similar.

Figure 2, Panel A shows estimates of the marginal burden of one day of major illness from Equation (2) at each quintile in the distribution of household economic status. For a household in the middle quintile, a day of illness in the past year is associated with reductions in annual per capita wage income of Rs. 45 [95% CI −91 to −5] and Rs 20 [95% CI −8 to −35] in the harmonized VDSA and IHDS samples, respectively. The corresponding increases in per capita medical spending due to illness are Rs 120 [95% CI 75 to 166] and Rs 88 [95% CI 83 to 93]. (Appendix Table A2 also shows that marginal burden values averaged across quintiles are generally similar). Panel B then presents average burden results, multiplying the marginal burden estimates in Panel A by 50 days (the average number of major illness days in a year for an individual in the IHDS sample), and expressed as a proportion of total household spending.

**Figure 2:**
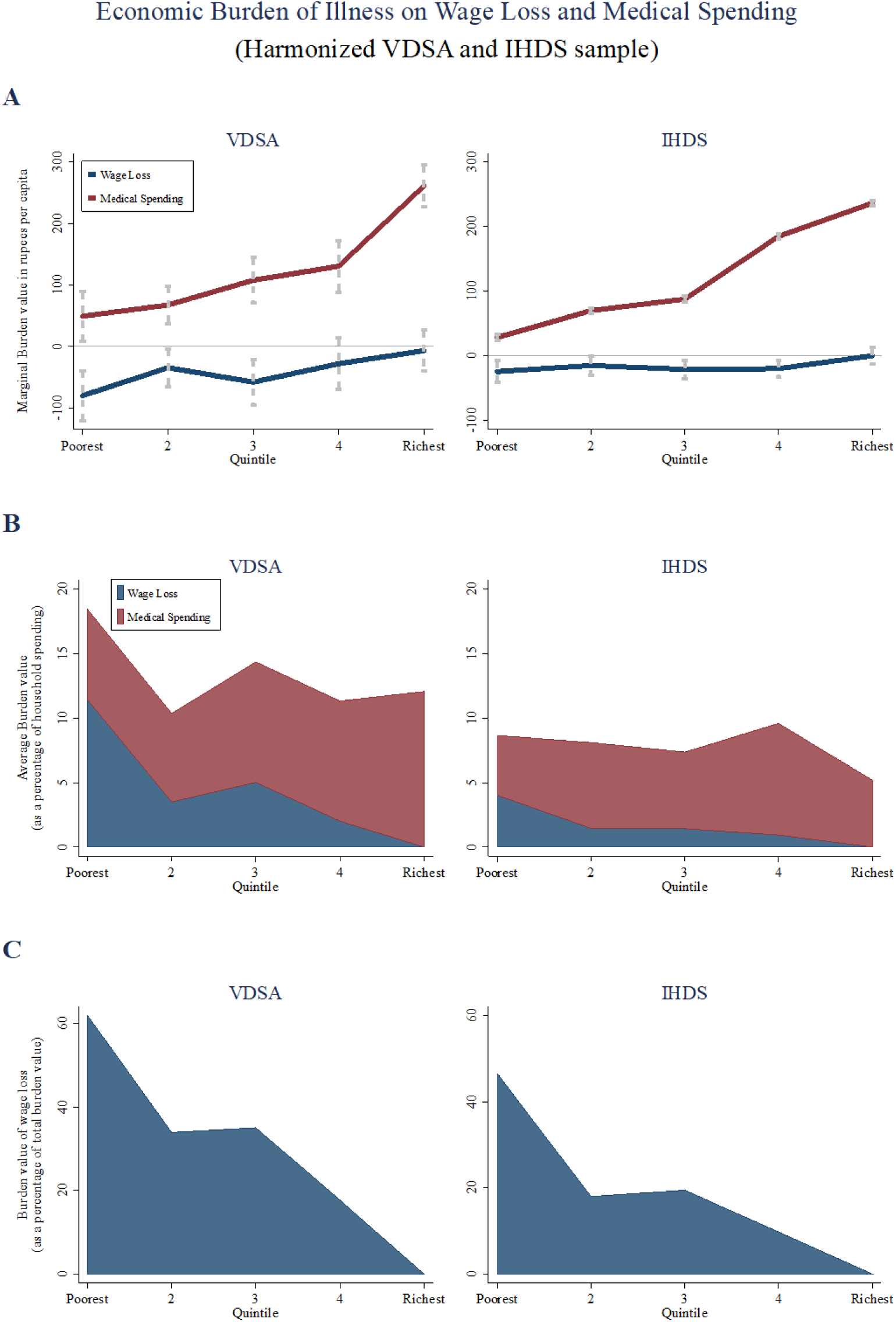
Panels A-D display comparable estimates from the harmonized VDSA (left) and IHDS (right) datasets. The IHDS contains data from 42,152 households in 2005 and again in 2011. Panel A presents estimates of the coefficients (from equation (2)) capturing the burden of illness on wage income and medical spending at different quantiles of the distribution of the household per capita spending. Both graphs include point-wise error bars showing the 95% confidence intervals for each coefficient. Panel B decomposes the total economic burden of illness into its two components – wage loss andmedical spending, and shows the economic burden as a percentage of total household spending. Panel C shows wage loss as a percentage of total household spending. In all panels, wage loss results are shown in blue, and medical spending results are shown in red.

Turning to how these results vary across the distribution of economic status, Figure 2, Panel A also shows that wage loss is generally greater among poorer households (in the lowest economic status quintile, for example) – although this magnitude is larger in the harmonized VDSA than in the IHDS sample. At the same time, in both data sets, the reduction in wage earnings due to illness is relatively smaller among those with higher economic status. Finally, in both the harmonized VDSA and IHDS samples, medical spending due to illness is relatively smaller at lower quintiles and larger at higher quintiles.

Figure 2, Panel C shows the share of the total burden of illness (wage loss and medical spending) due to lost wage earnings. Overall, the results – and the gradient in each outcome across the economic distribution – are generally consistent with those shown in Figure 1. And although this gradient is less steep in the IHDS sample than in the VDSA sample (in the bottom quintile, wage loss as a share of total burden is 47% in the IHDS sample, as opposed to 62% in the VDSA sample, for example), wage loss accounts for a substantially larger share of the total economic burden of illness among poorer households.

## Discussion

In this study, we show that wage loss accounts for a substantial portion of the total economic burden of illness in India. Importantly, this is disproportionately true among the poorest households. In the high-frequency VDSA sample, more than 80% of the economic burden of illness is due to wage loss among the poorest households – a figure that drops to about 20% among the most affluent households. Although this gradient across the economic distribution is somewhat less steep in the nationally-representative IHDS sample, the same general pattern is present. These findings suggest that even if successful, the current Universal Health Coverage (UHC) agenda may still not protect households in India, and potentially many low- and middle-income countries, from substantial financial risk due to illness.

Our paper makes three contributions to existing studies of the economic burden of illness. First, it directly estimates and compares the magnitude of wage loss and direct medical care expenses due to illness using detailed, high-frequency data on wage earnings (including informal activities), medical spending and illness. Past studies have largely focused on either total economic burden or medical care costs, but our direct comparison informs health policies focused on strengthening financial protection. Second, we show that the seemingly counterintuitive result that more affluent households face relatively greater financial risk due to illness (obtained using standard measures of financial risk, defined as household medical spending as a share of total spending) reported by other studies is reversed when wage loss is incorporated.^36,37^ Third, given the large number of longitudinal observations for each household in the VDSA sample, to the best of our knowledge, our paper is the first to produce *household-specific* estimates of the burden of illness, enabling us to trace-out both wage loss and medical care expenses flexibly across the distribution of household economic status – enabling a more granular analysis of disparities between the poor and more affluent.

Our study also has several limitations. First, in comparing the relative magnitude of economic burden due to wage loss versus medical care spending, it examines environments in which a degree of health insurance already exists. In India, for example, basic primary care provided by the public sector is in principle free, and many state health insurance schemes have emerged along with the national *Rashtriya Swasthya Bima Yojana* (RSBY) program. However, the majority of outpatient services in India are provided by private providers (frequently in the informal sector), and by 2014 (several years after the 2011 IHDS wave), only about one-fifth of hospitalized households had some form of formal insurance coverage.^41^ Second, the relationship between illness and income is complex and likely bi-directional (for instance, lower income and the concomitant poorer nutrition and other amenities may cause or exacerbate illness). Our approach, including fixed-effect analysis is a useful approach to addressing some of the problems posed by examining simple correlations between illness and income, but more work is clearly needed. Third, results from India, even those estimated in a nationally-representative sample, do not necessarily generalize to other countries. Nonetheless, India is the second most populous country in the world and is therefore an important marker of progress toward achieving Universal Health Coverage and the Sustainable Development Goals (SDGs).^42^

If there were greater policy emphasis placed on wage loss due to illness – and insurance against this risk (through wage loss or short-term disability insurance, for example), verification would present a difficult challenge in the design and implementation of such programs. In high-income countries, clinician certification of illness or disability is commonly required to initiate the provision of benefits. However, in environments like India, where a substantial share of healthcare providers are in the informal sector (with little or no formal clinical training and limited regulation),^36^ the challenges of verifying illness are likely to be more substantial. An imperfect solution to this challenge has been the emergence of hospital cash benefits or hospital confinement indemnity insurance, which provide benefits commonly indexed to a beneficiaries’ wage rate – but requiring hospitalization to initiate the payment of benefits (and hence may reflect inequality in access to health care, for example).

Moreover, we note that a considerable amount of informal insurance exists in low- and middle-income countries (through transfers from extended family members and social network members, for example). However, a large body of research shows that “consumption smoothing” through informal insurance mechanisms is both incomplete and inefficient in India and other countries,^31,43–45^ suggesting that potentially large welfare gains could still be achieved by the provision of formal wage loss or disability insurance, even to those with other sources of informal support. Further research should consider the types of illnesses (major morbidities versus short-term morbidities, for example) and other types of conditions (locations, occupations, household member relationships to the primary income earner, etc.) for which disability risk is most substantial.

Finally, there has been recent debate about how widely to interpret the aim of providing “financial protection” through UHC, with some arguing for a narrower focus on out-of-pocket medical spending alone,^46^ and others proposing a more expansive approach that incorporates wage loss due to illness.^47^ We inform this debate by providing direct quantitative evidence on the relative importance of both components of the burden of illness. In particular, we show that among the poorest households, wage loss is a much larger part of the full financial burden of illness – a result with important implications for the recently created Lancet Citizen’s Commission on Reimagining India’s Health System.^48^

## Data Availability

The de-identified VDSA and IHDS data used in this study are publicly available online:
VDSA: http://vdsa.icrisat.ac.in/
IHDS: http://www.icpsr.umich.edu/

http://www.icpsr.umich.edu/

## Author Contributions

GM and AS conceived the research idea. SJ, AS, GM, and AM contributed to the analytic design, data analysis, and interpretation. All authors contributed to the drafting and revising of the manuscript.

## Role of the Funding Organization or Sponsor

None

## APPENDIX

**TABLE A1.**
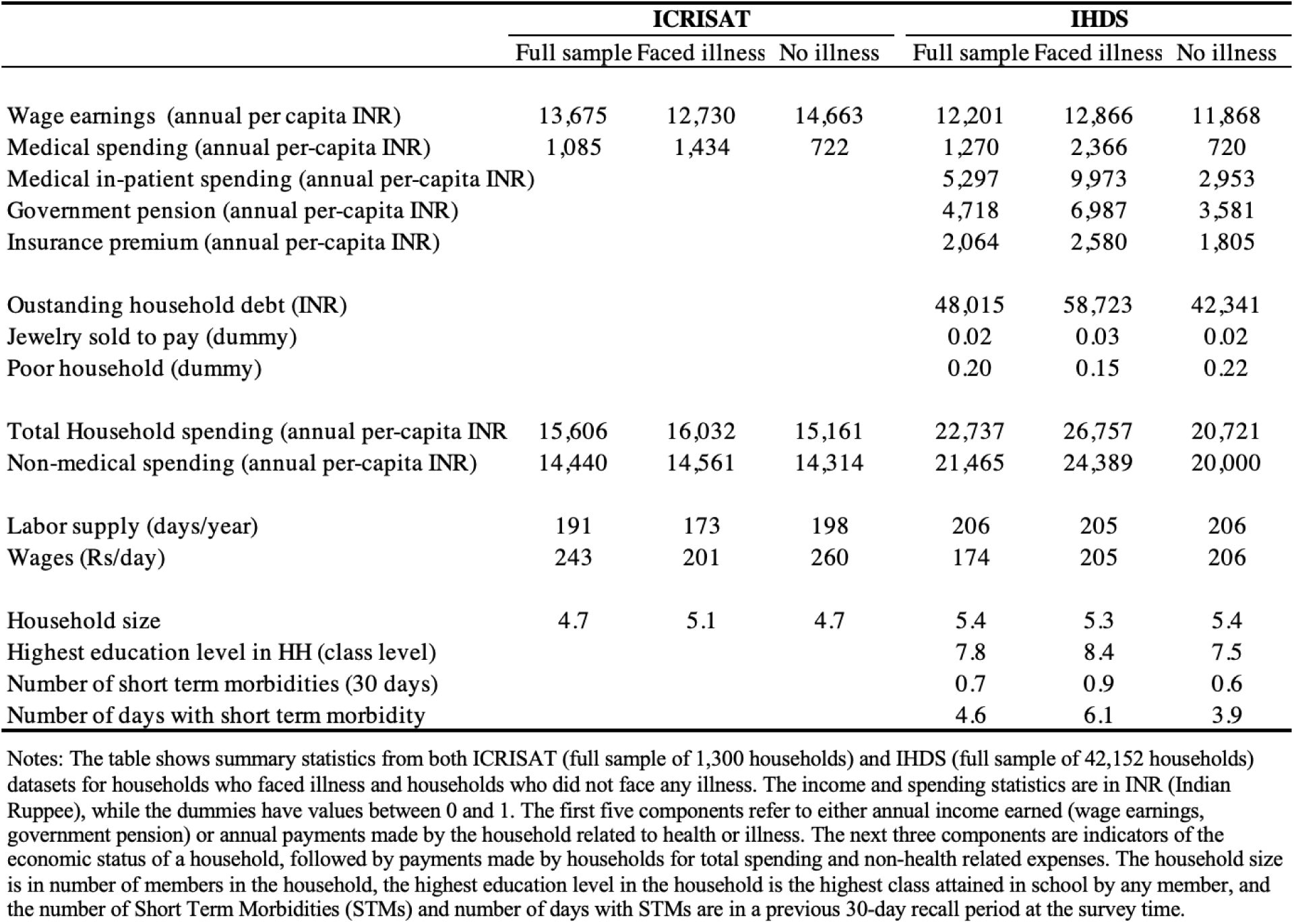
Summary statistics

| Table A1 : Summary statistics |  |  |  |  |  |  |
| --- | --- | --- | --- | --- | --- | --- |
|  | ICRISAT |  |  | IHDS |  |  |
|  | Full sample | Faced illness | No illness | Full sample | Faced illness | No illness |
| Wage earnings (annual per capita INR) | 13,675 | 12,730 | 14,663 | 12,201 | 12,866 | 11,868 |
| Medical spending (annual per-capita INR) | 1,085 | 1,434 | 722 | 1,270 | 2,366 | 720 |
| Medical in-patient spending (annual per-capita INR) |  |  |  | 5,297 | 9,973 | 2,953 |
| Government pension (annual per-capita INR) |  |  |  | 4,718 | 6,987 | 3,581 |
| Insurance premium (annual per-capita INR) |  |  |  | 2,064 | 2,580 | 1,805 |
| Oustanding household debt (INR) |  |  |  | 48,015 | 58,723 | 42,341 |
| Jewelry sold to pay (dummy) |  |  |  | 0.02 | 0.03 | 0.02 |
| Poor household (dummy) |  |  |  | 0.20 | 0.15 | 0.22 |
| Total Household spending (annual per-capita INR) | 15,606 | 16,032 | 15,161 | 22,737 | 26,757 | 20,721 |
| Non-medical spending (annual per-capita INR) | 14,440 | 14,561 | 14,314 | 21,465 | 24,389 | 20,000 |
| Labor supply (days/year) | 191 | 173 | 198 | 206 | 205 | 206 |
| Wages (Rs/day) | 243 | 201 | 260 | 174 | 205 | 206 |
| Household size | 4.7 | 5.1 | 4.7 | 5.4 | 5.3 | 5.4 |
| Highest education level in HH (class level) |  |  |  | 7.8 | 8.4 | 7.5 |
| Number of short term morbidities (30 days) |  |  |  | 0.7 | 0.9 | 0.6 |
| Number of days with short term morbidity |  |  |  | 4.6 | 6.1 | 3.9 |

Notes: The table shows summary statistics from both ICRISAT (full sample of 1,300 households) and IHDS (full sample of 42,152 households) datasets for households who faced illness and households who did not face any illness. The income and spending statistics are in INR (Indian Ruppee), while the dummies have values between 0 and 1. The first five components refer to either annual income earned (wage earnings, government pension) or annual payments made by the household related to health or illness. The next three components are indicators of the economic status of a household, followed by payments made by households for total spending and non-health related expenses. The household size is in number of members in the household, the highest education level in the household is the highest class attained in school by any member, and the number of Short Term Morbidities (STMs) and number of days with STMs are in a previous 30-day recall period at the survey time.

**TABLE A2.** Regression results for the economic burden of illness

| Table A2: Regression results for the economic burden of illness |  |  |
| --- | --- | --- |
|  | Medical spending | Wage Income |
| ***VDSA sample (monthly per capita)*** |  |  |
| Illness (Did any individual face illness, Yes = 1) | 126.035***<br>(8.189) | -77.685***<br>(10.662) |
| Observations | 79493 | 79513 |
| ***Harmonized VDSA sample (annual per capita)*** |  |  |
| Illness (# of ill days) | 126.274***<br>(8.368) | -38.756***<br>(8.507) |
| Observations | 28433 | 28433 |
| ***IHDS (annual per capita)*** |  |  |
| Illness (# of ill days) | 135.227***<br>(1.100) | -17.549***<br>(3.477) |
| Observations | 299992 | 299992 |

Notes: This table reports the average burden of illness on medical spending and wage income. For the *VDSA sample (monthly per capita)*, the estimating equation is similar to equation (1), expect that the illness variable is common across households (not household-specific) and captures the average burden of illness. The unit of observation is household-month, the estimation includes household and month fixed effects, illness is an indicator variable equal to 1 if any member in the household reported an illness, and 0 otherwise. For the *Harmonized VDSA sample (annual per capita)* and *IHDS (annual per capita)*, the estimating equation is similar to equation (2), expect that the illness variable is common across households (without the quintile interaction). The unit of observation is individual-year, the estimation includes individual and year fixed effects, and illness variable is the number of days of illness in the previous year. Medical spending and wage income are measured in rupees per capita. \* p<0.10 \*\*p<0.05 \*\*\* p<0.01

**Figure A1:**
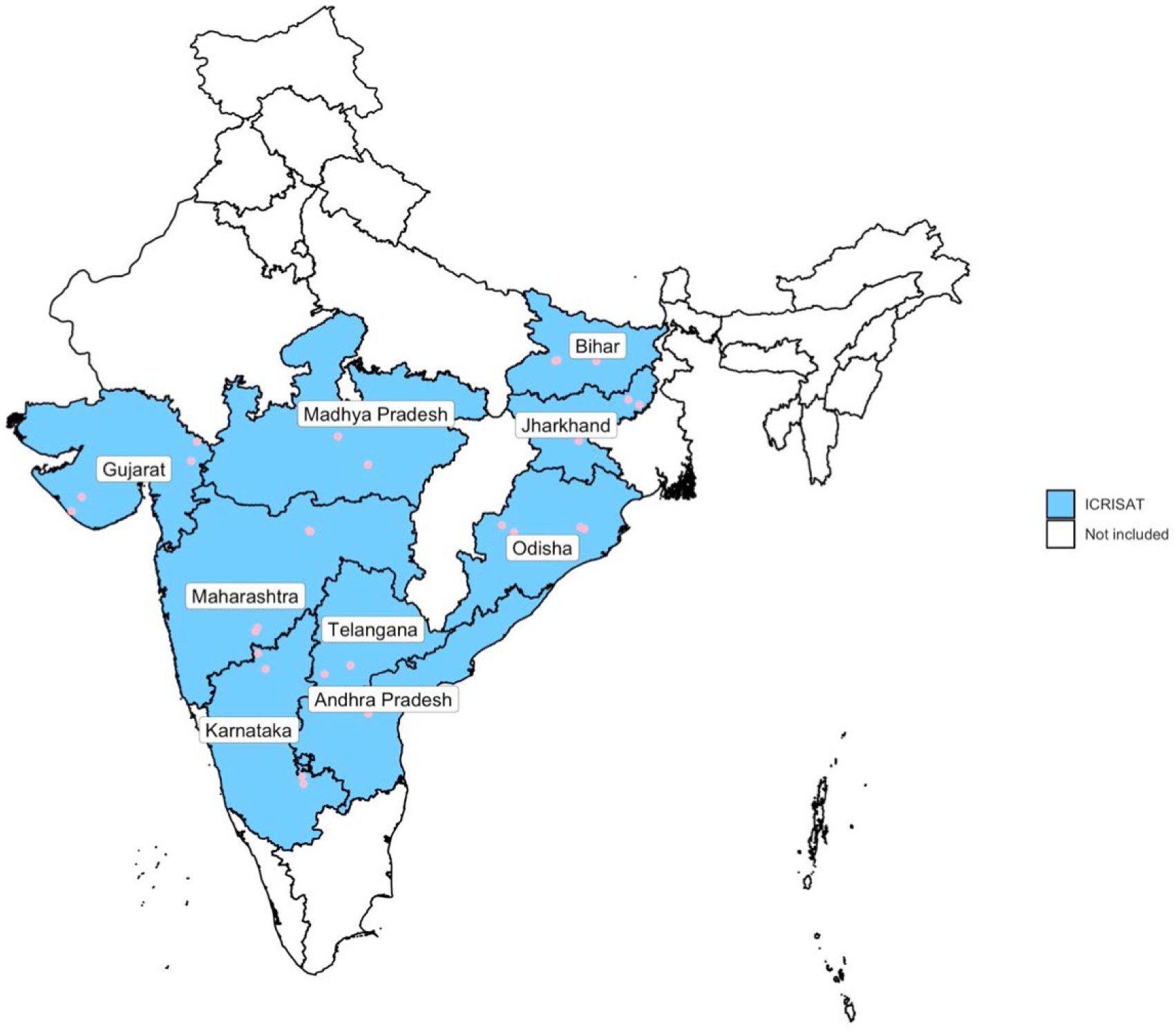
Locations of ICRISAT VDSA villages The map of India shows the locations of the villages in the ICRSAT dataset across nine states

